# Smaller Targets and a Shorter Warning: Changes in the Cyberthreat to US Health Care Providers Before and After Generative AI

**DOI:** 10.64898/2026.11.17.26368300

**Authors:** Ryutaro Nakagawa, Yuya Yasuhara, Akiyo Shimoyama, Yuki Nishi, Yuki Kubota

**Author notes:** Corresponding author: Ryutaro Nakagawa, Centralmedience Co., Ltd., SOUTH PORT Shinagawa 8F, 2-12-32 Konan, Minato-ku, Tokyo 108-0075, Japan. This article is a preprint and has not been peer reviewed. This is version 3. Version 1 (November 2026) reported period comparisons. Version 2 made interrupted time-series regression the primary analysis and added an analysis of alternative breakpoints. This version adds a formal test for the existence of a break and a confidence set for its date, the minimum detectable change in trend behind the negative finding about generative artificial intelligence, two control series drawn from the same reporting system, sensitivity to the size threshold and to reporting maturity, a check on the pace at which the vulnerability catalog was maintained, and residual diagnostics. It applies the search over alternative breakpoints to every outcome rather than to breach size alone; as a result the claim that the change in exploitation speed predates generative artificial intelligence has been withdrawn, because December 2022 falls inside the confidence set for that series. Breach data were re-downloaded on November 28, 2026. The processed data and the analysis code are attached as a supplementary file.

## Abstract

Hospitals are critical infrastructure, and attacks on them harm patients at the facility attacked and at its neighbors. Two assumptions shape where defensive investment and national support go: that large organizations are the main targets, and that generative AI is accelerating the threat. We tested both with two public US sources, using interrupted time-series regression with a break at December 2022, the month after ChatGPT was released. We used 578 entries from the federal catalog of known exploited vulnerabilities, published between November 2021 and October 2025, and 3,603 provider hacking incidents among 8,093 federal breach reports. Monthly incidents rose 32.2% per year before the break and did not accelerate afterward; an acceleration of 21% per year would have been detected. The share of incidents affecting fewer than 5,000 individuals fell before the break and rose afterward. A break exists (bootstrap P = .006), but it is dated August to November 2021, holds at every size threshold, and is absent from two control series in the same portal, so it is not an artifact of reporting. The share of vulnerabilities exploited within 7 days rose and then flattened; for that series alone the timing cannot be separated from the arrival of generative AI, but the change is a deceleration, not the anticipated acceleration. Nothing accelerated after generative AI became available. Exploitation now follows disclosure within a week while remediation takes 2 months. Reducing what is reachable from the internet, and sustaining care without electronic systems, are the actionable priorities.

## Introduction

Cyberattacks on hospitals are a health security problem rather than only an information technology problem.^1^ In the United States, in-hospital mortality among patients who were already admitted when an attack began rose by about a third.^2^ An attack on one health system brought more emergency visits, ambulance arrivals, stroke codes, and cardiac arrests to neighboring hospitals that were never attacked,^3–5^ so the consequences of a single compromised organization are regional. Health systems elsewhere have had care disrupted for weeks to months.^6^ Protecting health care delivery from these events belongs with the other hazards that preparedness planning addresses.

Current thinking about that hazard rests on two observations. Between 2016 and 2021, ransomware attacks became concentrated among larger organizations: the share of attacks on organizations with multiple facilities rose from 41.9% to 76.9%,^7^ and hospitals that were attacked had higher revenues and were less often rural than those that were not.^8^ At the same time, the interval between public disclosure of a software vulnerability and its exploitation has been shrinking,^9,10^ and national authorities expect generative artificial intelligence to increase both the volume and the impact of cyberattacks.^11^ The resulting picture is one in which large organizations are the main targets and artificial intelligence is the next accelerant. Both halves of that picture shape where defensive investment goes, which organizations national programs are designed around, and what preparedness planners are told to expect.

Neither half has been tested against data extending past the point at which generative artificial intelligence became widely available. Peer-reviewed studies of ransomware in health care end in 2021.^7,8^ Analyses of exploitation timelines come mostly from industry, report descriptive medians, and do not ask when the shortening began.^9,10,12^ Recent work using news-derived event data has mapped the healthcare threat landscape and shown that reported attacks grew substantially between 2017 and 2023, but it describes volume and attack type rather than testing when the shape of the threat changed.^13^

The distinction matters for planning. If these changes did not accelerate after November 2022, the emphasis on artificial intelligence as the proximate driver is misplaced, and the more mundane question of what remains reachable from the internet becomes the operative one. It also determines whether national support should still be designed around large health systems. We therefore asked three questions. Did the time from disclosure of a vulnerability to its confirmed exploitation shorten, and did that shortening accelerate after generative artificial intelligence became available? Did hacking breaches at US health care providers change in size, and in which direction? And is the arrival of that technology the point at which either series changed, compared with other candidate dates?

## Methods

### Design and data sources

This retrospective observational study used only publicly available data. No individual-level health information was accessed and no human participants were involved, so institutional review board approval was not required.

**Exploitation timelines.** We downloaded the catalog of known exploited vulnerabilities maintained by the US Cybersecurity and Infrastructure Security Agency (version 2026.10.16; 1,748 entries),^14^ which lists vulnerabilities with reliable evidence of exploitation in the wild, and took each vulnerability’s publication date from the public CVE List V5 repository.^15^ Time to confirmed exploitation was the date of addition to the catalog minus the publication date, a measure also used in industry reporting.^12^ The catalog launched on November 3, 2021 with a backlog, so we excluded vulnerabilities published before that date. Recent vulnerabilities have had less time to be added, so we counted only additions within 365 days of publication and dropped publication months with less follow-up, leaving vulnerabilities published from November 2021 through October 2025. Edge devices — virtual private network appliances, firewalls, and remote-access gateways — were identified from an explicit vendor and product list, given in the supplementary material.

**Breach size.** On November 28, 2026 we downloaded all reports from the breach portal of the Office for Civil Rights of the US Department of Health and Human Services, which lists breaches of unsecured protected health information affecting 500 or more individuals. After removing duplicates, 8,093 reports remained. We kept reports filed by health care providers whose breach type was exactly “Hacking/IT Incident” (n = 3,603). For each month we counted incidents, those affecting fewer than 5,000 individuals, and those listing email as a location of breached information. The number of individuals affected is the only size-related field in the portal; we used the share of incidents affecting fewer than 5,000 individuals as a proxy for smaller events, as in earlier work on this source.^7^

### Statistical analysis

The primary analysis was an interrupted time-series regression. For the monthly count of provider hacking incidents we fitted a negative binomial regression with a linear time term, an indicator for the post-break period, an interaction between time and that indicator, and sine and cosine terms at annual and semi-annual frequencies to absorb seasonality in reporting. For proportions — the share of incidents affecting fewer than 5,000 individuals, and the shares of vulnerabilities exploited within 7 days and on or before disclosure — we fitted binomial regressions with the same structure, omitting seasonal terms for the vulnerability series. Standard errors were Newey–West with 12 lags for the monthly breach series and 6 for the vulnerability series. Estimates are rate ratios or odds ratios per year with 95% confidence intervals. The interaction term is the ratio of the post-break trend to the pre-break trend, so 1 means no change.

The break was set at December 2022, the first full month after the public release of ChatGPT on November 30, 2022. The primary breach window was January 2016 to December 2025; 2026 was excluded because reports are still accruing, and a sensitivity analysis extended the window to June 2026.

A single pre-specified break can fit well even when the true change lies elsewhere, so we refitted each model with every month from January 2019 to December 2024 as the break and ranked the 72 candidates by the Akaike information criterion. As descriptive support, exploitation times were compared between periods with the Mann–Whitney U test, across the six half-years of 2023 to 2025 with the Kruskal–Wallis test, and linear trends in annual proportions with the Cochran–Armitage test. Analyses were done in Python 3.11 with statsmodels 0.14 and SciPy.

### Testing the break, and testing the result against alternatives

Ranking candidate breakpoints shows which date fits best, not whether any break exists. We therefore tested the null hypothesis of no break anywhere in the window, using the largest likelihood-ratio statistic over the 72 candidate months against a model with trend and seasonality but no break. Its null distribution came from a parametric bootstrap with 999 replications, drawing monthly shares from a beta-binomial distribution matched to the fitted values and the observed overdispersion, so that simulated series are as noisy as the real one. We treat candidate months within 2 units of the best Akaike information criterion as a confidence set for the break date, and applied the same search to every outcome.

For the question about generative artificial intelligence the result is a negative one, so the relevant quantity is not only the estimate but what size of effect the study could have detected. We report the minimum change in trend that would have been detected with 80% power at a two-sided 5% level, given the standard errors obtained.

Further checks address the most likely alternative explanations. A change in reporting rather than in attacks would affect other reports in the same portal, so we fitted the same models to two control series from that system: breaches reported by health care providers whose type was not “Hacking/IT Incident”, and hacking incidents reported by covered entities that are not health care providers. The 5,000-individual cutoff is arbitrary, so we repeated the analysis at 1,000, 2,500 and 10,000, and because recent reports are still accruing we also ended the series in June 2025 and December 2024. Quick exploitation could reflect how actively the catalog was maintained rather than how fast attackers moved, so we refitted that model with the logarithm of catalog additions in the month as a covariate and dropped the catalog’s first 3 and 6 months. We also checked residual autocorrelation and the choice of Newey–West lag.

### Responsible reporting of vulnerability information

Every vulnerability analyzed here was already public and already confirmed as exploited before it entered the federal catalog. We report aggregate timing and device categories only. No exploitation method, configuration detail, or information identifying an exposed organization appears in this article or its supplementary material, so the analysis adds nothing to an attacker’s capability while giving defenders the timing they need.

## Results

### Exploitation timelines

Of 1,748 catalog entries, 578 met the inclusion criteria. Median time to confirmed exploitation fell from 24 days (interquartile range 3 to 106) for vulnerabilities published in the second half of 2021 and first half of 2022, to 0 days (−4 to 50) for the second half of 2022, and was 6 days (0 to 49) across 2023 to 2025 (Table 1, Figure 1). Exploitation took longer in the first period than later (Mann–Whitney P = .007). Across the six half-years of 2023 to 2025 there was no difference (Kruskal–Wallis P = .40). The share exploited on or before the publication date was 20.8%, 54.9%, and 27.3% in the three periods. For edge devices (n = 85) the median was 5 days and 71.8% were exploited within 30 days.

**Figure 1.**
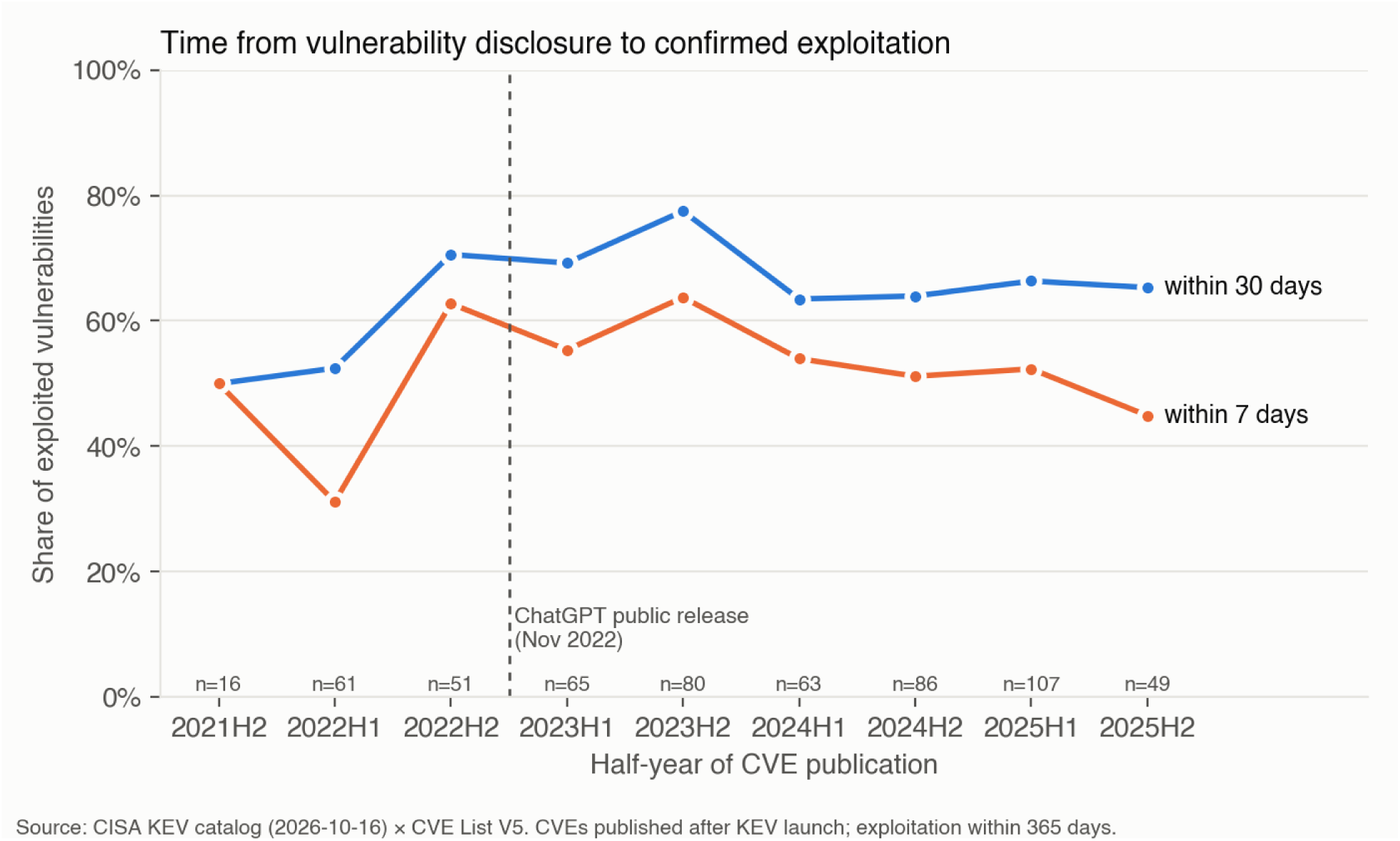
Median time from disclosure to confirmed exploitation, by half-year of publication.

**TABLE 1.** Time from disclosure to confirmed exploitation, by half-year of CVE publication.

| **Half-year of publication** | **Vulnerabilities, No.** | **Median days to exploitation (IQR)** | **Exploited on or before disclosure, %** | **Within 7 days, %** | **Within 30 days, %** |
|---|---|---|---|---|---|
| 2021H2 | 16 | 20 (0 to 143) | 31.2 | 50.0 | 50.0 |
| 2022H1 | 61 | 24 (4 to 100) | 18.0 | 31.1 | 52.5 |
| 2022H2 | 51 | 0 (-4 to 50) | 54.9 | 62.7 | 70.6 |
| 2023H1 | 65 | 4 (0 to 88) | 33.8 | 55.4 | 69.2 |
| 2023H2 | 80 | 4 (0 to 14) | 30.0 | 63.7 | 77.5 |
| 2024H1 | 63 | 6 (0 to 102) | 28.6 | 54.0 | 63.5 |
| 2024H2 | 86 | 7 (1 to 62) | 24.4 | 51.2 | 64.0 |
| 2025H1 | 107 | 6 (0 to 39) | 25.2 | 52.3 | 66.4 |
| 2025H2 | 49 | 10 (1 to 48) | 22.4 | 44.9 | 65.3 |
| Pooled, 2021H2 to 2022H1 | 77 | 24 (3 to 106) | 20.8 | 35.1 | 51.9 |
| Pooled, 2022H2 | 51 | 0 (-4 to 50) | 54.9 | 62.7 | 70.6 |
| Pooled, 2023H1 to 2025H2 | 450 | 6 (0 to 49) | 27.3 | 54.0 | 67.8 |
| Edge devices, all periods | 85 | 5 (0 to 35) | 29.4 | 55.3 | 71.8 |

Note: Mann-Whitney 2021H2-2022H1 vs later: U=22610 P=0.0073
Note: Kruskal-Wallis across 6 half-years 2023H1-2025H2: H=5.16 P=0.40

In the interrupted time-series models, the share exploited within 7 days rose steeply before December 2022 (odds ratio 5.50 per year, 95% confidence interval 2.13 to 14.16) and the trend reversed afterward (ratio of trends 0.144, 0.055 to 0.374, P < .001), with no level change at the break itself (0.800, 0.494 to 1.295, P = .36) (Table 3). The share exploited on or before disclosure behaved the same way; its level fell at the break (0.368, 0.234 to 0.581, P < .001) because the peak occurred in the months immediately before it.

**TABLE 2.** Hacking/IT incidents reported by US health care providers, by year of report.

| **Year of report** | **Incidents, No.** | **Median individuals affected** | **Affecting <5,000 individuals, %** | **Email listed as location, %** |
|---|---|---|---|---|
| 2016 | 100 | 8,750 | 42.0 | 17.0 |
| 2017 | 123 | 6,642 | 43.1 | 32.5 |
| 2018 | 124 | 6,098 | 46.0 | 51.6 |
| 2019 | 251 | 7,678 | 40.6 | 53.8 |
| 2020 | 363 | 9,058 | 39.1 | 39.9 |
| 2021 | 386 | 8,862 | 39.9 | 31.6 |
| 2022 | 415 | 14,203 | 31.8 | 30.6 |
| 2023 | 373 | 12,317 | 31.4 | 21.4 |
| 2024 | 452 | 10,681 | 39.4 | 26.1 |
| 2025 | 506 | 6,418 | 45.3 | 24.9 |
| 2026 | 421 | 5,086 | 49.4 | 22.6 |

Note: Cochran-Armitage trend in share <5,000, 2016-2022: z=-2.90 P=0.0037
Note: Cochran-Armitage trend in share <5,000, 2022-2025: z=4.77 P=1.8e-06
Note: 2026 is partial: reports submitted through October 2026. Portal retrieved November 28, 2026.

**TABLE 3.** Interrupted time-series estimates (break at December 2022, the month after the public release of ChatGPT)

| **Outcome** | **Term** | **Estimate (95% CI)** | **P** |
|---|---|---|---|
| Monthly number of hacking/IT incidents (rate ratio) | Underlying trend, per year | 1.322 (1.262–1.385) | <0.001 |
| Monthly number of hacking/IT incidents (rate ratio) | Level change at December 2022 | 0.601 (0.488–0.741) | <0.001 |
| Monthly number of hacking/IT incidents (rate ratio) | Change in trend after December 2022, per year | 0.883 (0.771–1.012) | 0.073 |
| Share of incidents affecting <5000 individuals (odds ratio) | Underlying trend, per year | 0.923 (0.866–0.983) | 0.013 |
| Share of incidents affecting <5000 individuals (odds ratio) | Level change at December 2022 | 0.809 (0.601–1.089) | 0.163 |
| Share of incidents affecting <5000 individuals (odds ratio) | Change in trend after December 2022, per year | 1.409 (1.285–1.545) | <0.001 |
| Share of vulnerabilities exploited within 7 days (odds ratio) | Underlying trend, per year | 5.497 (2.134–14.159) | <0.001 |
| Share of vulnerabilities exploited within 7 days (odds ratio) | Level change at December 2022 | 0.800 (0.494–1.295) | 0.363 |
| Share of vulnerabilities exploited within 7 days (odds ratio) | Change in trend after December 2022, per year | 0.144 (0.055–0.374) | <0.001 |
| Share exploited on or before disclosure (odds ratio) | Underlying trend, per year | 8.753 (3.435–22.303) | <0.001 |
| Share exploited on or before disclosure (odds ratio) | Level change at December 2022 | 0.368 (0.234–0.581) | <0.001 |
| Share exploited on or before disclosure (odds ratio) | Change in trend after December 2022, per year | 0.085 (0.033–0.222) | <0.001 |

Note: Counts: negative binomial regression; shares: binomial regression. All models include a linear time term and Newey-West standard errors (12 lags for monthly breach series, 6 for vulnerability series). Breach models include seasonal terms.

### Breach size among health care providers

Among 3,603 provider hacking incidents, the share affecting fewer than 5,000 individuals declined from 42.0% in 2016 to 31.8% in 2022 (Cochran–Armitage P = .004), then rose to 45.3% in 2025 (2022 to 2025 P < .001) and stood at 49.4% in the partial year 2026 (Table 2, Figure 2). The median number of individuals affected peaked at 14,203 in 2022 and fell to 6,418 in 2025 and 5,086 in 2026. Email appeared as a location of breached information in 53.8% of incidents in 2019 and in 21% to 26% from 2023 onward.

**Figure 2.**
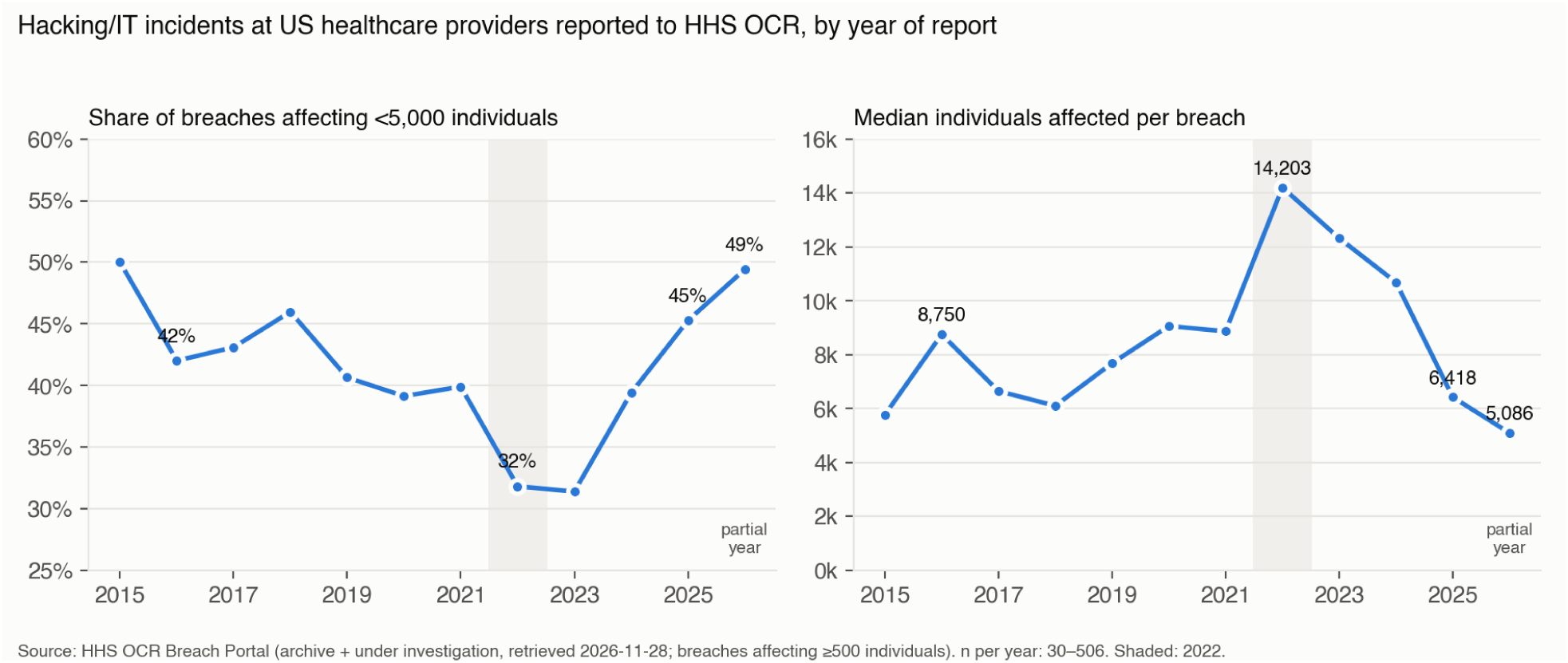
Share of provider hacking incidents affecting fewer than 5,000 individuals, and median individuals affected, by year.

Over the 120 months from January 2016 to December 2025 (3,093 incidents), the monthly count rose 32.2% per year before the break (rate ratio 1.322, 95% confidence interval 1.262 to 1.385, P < .001). It fell at the break (0.601, 0.488 to 0.741, P < .001) and the trend did not accelerate afterward; if anything it slowed (0.883, 0.771 to 1.012, P = .07). The share affecting fewer than 5,000 individuals fell before the break (0.923 per year, 0.866 to 0.983, P = .01) and rose afterward (1.409, 1.285 to 1.545, P < .001), with no level change at the break (0.809, 0.601 to 1.089, P = .16). Extending the window to June 2026 changed nothing (supplementary material).

### When did the change happen?

The null hypothesis of no break anywhere between January 2019 and December 2024 was rejected (largest likelihood-ratio statistic 34.3 at August 2021; bootstrap P = .006), so a break exists in breach size. It is not where generative artificial intelligence arrived. The best-fitting month was August 2021, and the confidence set — candidate months within 2 Akaike information criterion units of the best — was August to November 2021. December 2022 lay outside it, ranking 28th of 72 and fitting about 200 times less well than the best month (Figure 3). The reversal in breach size therefore began about a year before generative artificial intelligence became publicly available.

**Figure 3.**
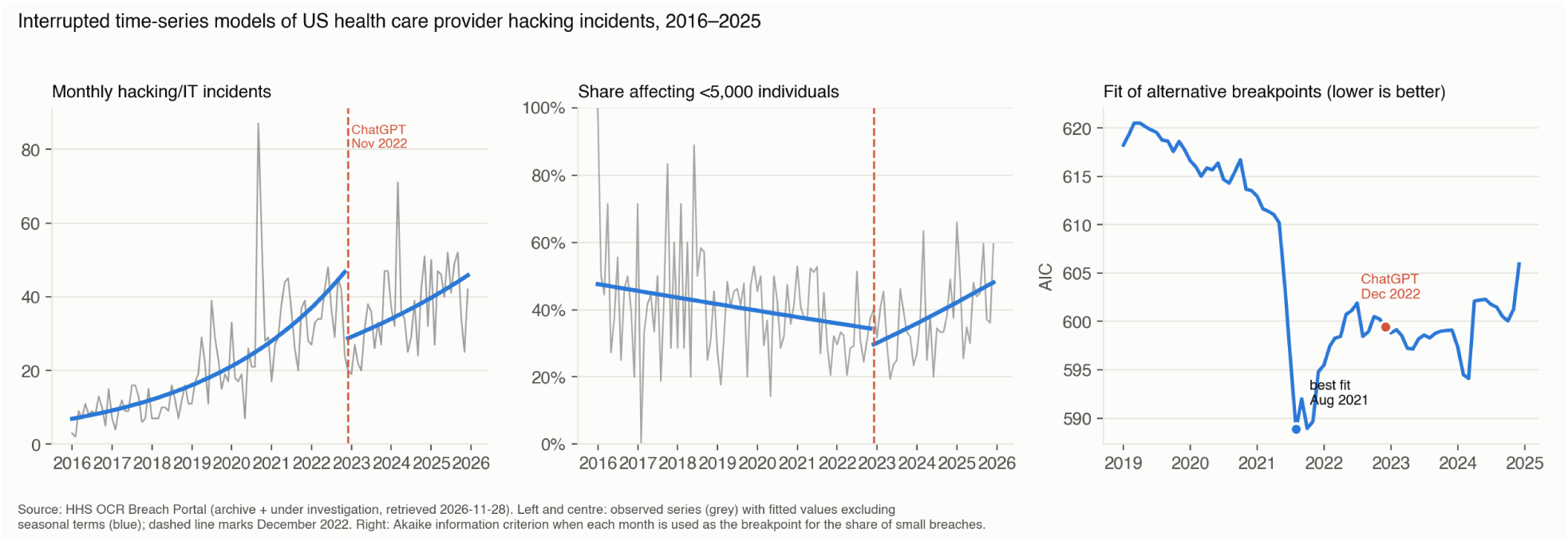
Interrupted time-series fits and model fit across all candidate breakpoints.

The same search separates the other outcomes (Table 4). For the monthly count the best-fitting break was September 2020, with December 2022 ranked 46th of 72, well outside the confidence set. For the share of vulnerabilities exploited within 7 days, however, the best-fitting break was May 2022 and December 2022 fell inside the confidence set, which ran from May 2022 to March 2023; for the share exploited on or before disclosure the best fit was January 2023, with December 2022 ranked third of 36. For the vulnerability series the timing of the change therefore cannot be separated from the arrival of generative artificial intelligence. Its direction can: the change is a flattening, not an acceleration.

**TABLE 4.**
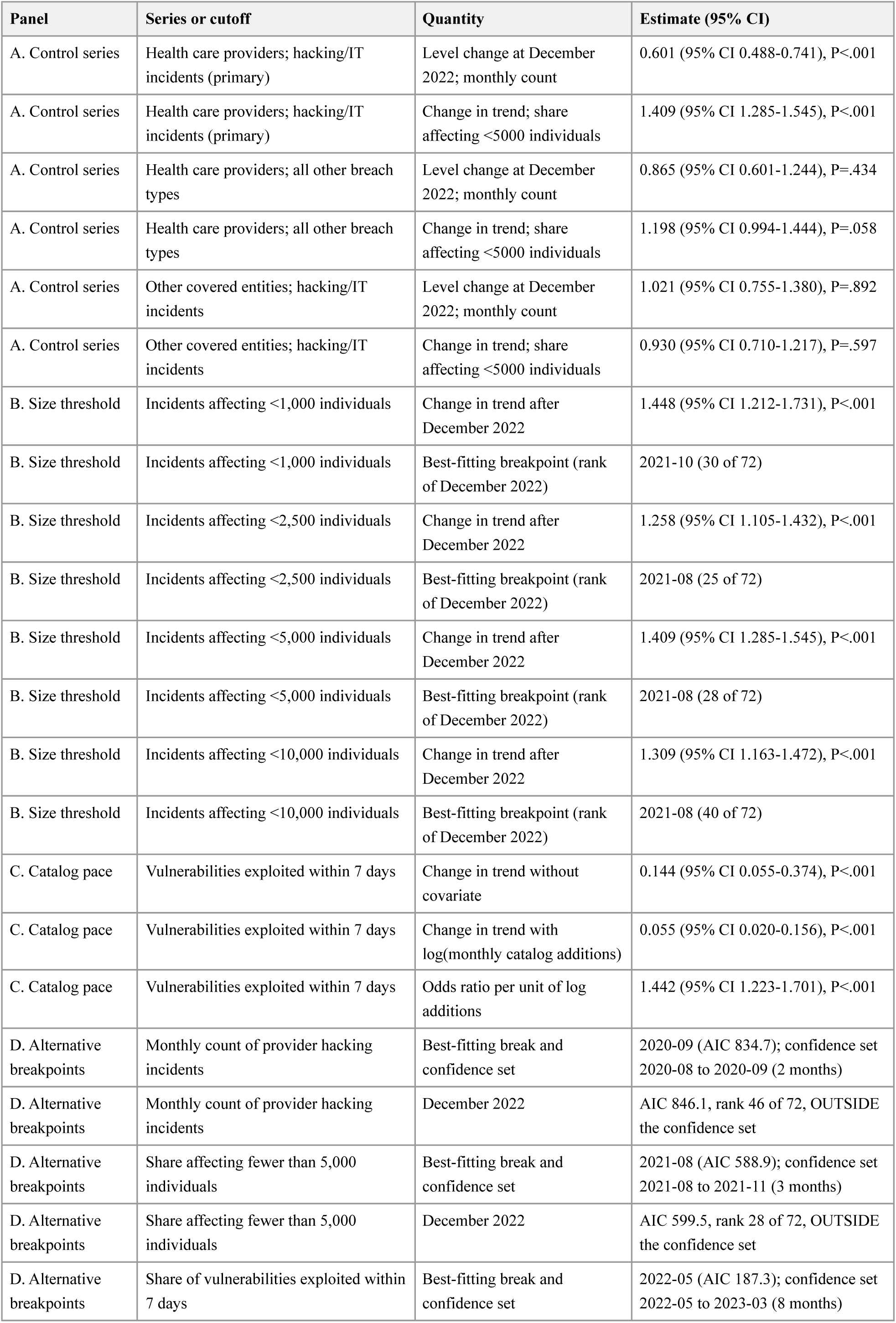
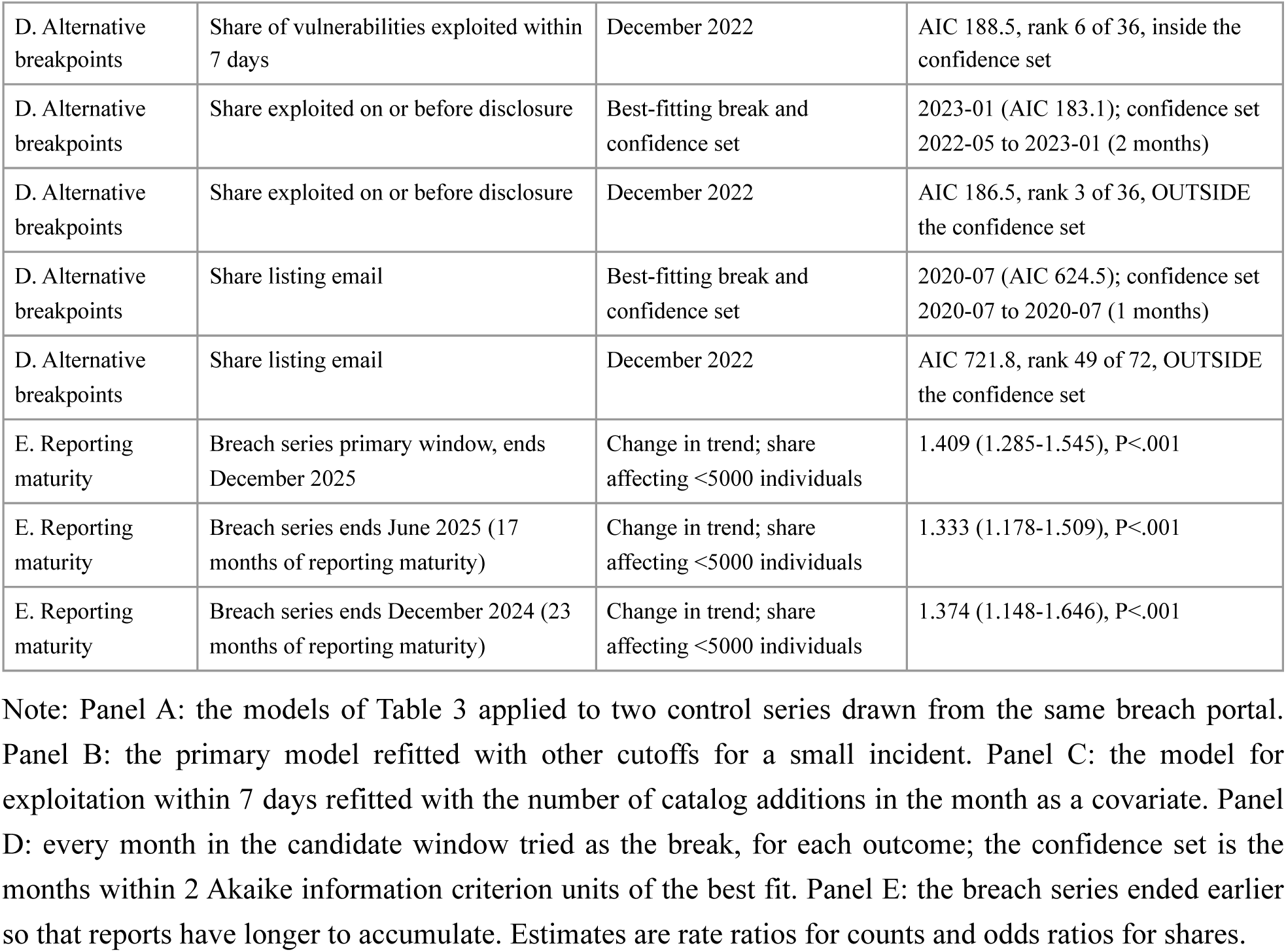
Robustness checks.

| Panel | Series or cutoff | Quantity | Estimate (95% CI) |
|---|---|---|---|
| A. Control series | Health care providers; hacking/IT incidents (primary) | Level change at December 2022; monthly count | 0.601 (95% CI 0.488-0.741), P<.001 |
| A. Control series | Health care providers; hacking/IT incidents (primary) | Change in trend; share affecting <5000 individuals | 1.409 (95% CI 1.285-1.545), P<.001 |
| A. Control series | Health care providers; all other breach types | Level change at December 2022; monthly count | 0.865 (95% CI 0.601-1.244), P=.434 |
| A. Control series | Health care providers; all other breach types | Change in trend; share affecting <5000 individuals | 1.198 (95% CI 0.994-1.444), P=.058 |
| A. Control series | Other covered entities; hacking/IT incidents | Level change at December 2022; monthly count | 1.021 (95% CI 0.755-1.380), P=.892 |
| A. Control series | Other covered entities; hacking/IT incidents | Change in trend; share affecting <5000 individuals | 0.930 (95% CI 0.710-1.217), P=.597 |
| B. Size threshold | Incidents affecting <1,000 individuals | Change in trend after December 2022 | 1.448 (95% CI 1.212-1.731), P<.001 |
| B. Size threshold | Incidents affecting <1,000 individuals | Best-fitting breakpoint (rank of December 2022) | 2021-10 (30 of 72) |
| B. Size threshold | Incidents affecting <2,500 individuals | Change in trend after December 2022 | 1.258 (95% CI 1.105-1.432), P<.001 |
| B. Size threshold | Incidents affecting <2,500 individuals | Best-fitting breakpoint (rank of December 2022) | 2021-08 (25 of 72) |
| B. Size threshold | Incidents affecting <5,000 individuals | Change in trend after December 2022 | 1.409 (95% CI 1.285-1.545), P<.001 |
| B. Size threshold | Incidents affecting <5,000 individuals | Best-fitting breakpoint (rank of December 2022) | 2021-08 (28 of 72) |
| B. Size threshold | Incidents affecting <10,000 individuals | Change in trend after December 2022 | 1.309 (95% CI 1.163-1.472), P<.001 |
| B. Size threshold | Incidents affecting <10,000 individuals | Best-fitting breakpoint (rank of December 2022) | 2021-08 (40 of 72) |
| C. Catalog pace | Vulnerabilities exploited within 7 days | Change in trend without covariate | 0.144 (95% CI 0.055-0.374), P<.001 |
| C. Catalog pace | Vulnerabilities exploited within 7 days | Change in trend with log(monthly catalog additions) | 0.055 (95% CI 0.020-0.156), P<.001 |
| C. Catalog pace | Vulnerabilities exploited within 7 days | Odds ratio per unit of log additions | 1.442 (95% CI 1.223-1.701), P<.001 |
| D. Alternative breakpoints | Monthly count of provider hacking incidents | Best-fitting break and confidence set | 2020-09 (AIC 834.7); confidence set 2020-08 to 2020-09 (2 months) |
| D. Alternative breakpoints | Monthly count of provider hacking incidents | December 2022 | AIC 846.1, rank 46 of 72, OUTSIDE the confidence set |
| D. Alternative breakpoints | Share affecting fewer than 5,000 individuals | Best-fitting break and confidence set | 2021-08 (AIC 588.9); confidence set 2021-08 to 2021-11 (3 months) |
| D. Alternative breakpoints | Share affecting fewer than 5,000 individuals | December 2022 | AIC 599.5, rank 28 of 72, OUTSIDE the confidence set |
| D. Alternative breakpoints | Share of vulnerabilities exploited within 7 days | Best-fitting break and confidence set | 2022-05 (AIC 187.3); confidence set 2022-05 to 2023-03 (8 months) |
| D. Alternative breakpoints | Share of vulnerabilities exploited within 7 days | December 2022 | AIC 188.5, rank 6 of 36, inside the confidence set |
| D. Alternative breakpoints | Share exploited on or before disclosure | Best-fitting break and confidence set | 2023-01 (AIC 183.1); confidence set 2022-05 to 2023-01 (2 months) |
| D. Alternative breakpoints | Share exploited on or before disclosure | December 2022 | AIC 186.5, rank 3 of 36, OUTSIDE the confidence set |
| D. Alternative breakpoints | Share listing email | Best-fitting break and confidence set | 2020-07 (AIC 624.5); confidence set 2020-07 to 2020-07 (1 months) |
| D. Alternative breakpoints | Share listing email | December 2022 | AIC 721.8, rank 49 of 72, OUTSIDE the confidence set |
| E. Reporting maturity | Breach series primary window, ends December 2025 | Change in trend; share affecting <5000 individuals | 1.409 (1.285-1.545), P<.001 |
| E. Reporting maturity | Breach series ends June 2025 (17 months of reporting maturity) | Change in trend; share affecting <5000 individuals | 1.333 (1.178-1.509), P<.001 |
| E. Reporting maturity | Breach series ends December 2024 (23 months of reporting maturity) | Change in trend; share affecting <5000 individuals | 1.374 (1.148-1.646), P<.001 |

Note: Panel A: the models of Table 3 applied to two control series drawn from the same breach portal. Panel B: the primary model refitted with other cutoffs for a small incident. Panel C: the model for exploitation within 7 days refitted with the number of catalog additions in the month as a covariate. Panel D: every month in the candidate window tried as the break, for each outcome; the confidence set is the months within 2 Akaike information criterion units of the best fit. Panel E: the breach series ended earlier so that reports have longer to accumulate. Estimates are rate ratios for counts and odds ratios for shares.

### How large an acceleration could have been detected?

Because the finding about generative artificial intelligence is negative, we report what the study could have detected. For monthly counts, a change in trend of 1.214 or more — an acceleration of at least 21% per year on top of the existing trend — would have been detected with 80% power; the observed value was 0.883. For the share of small incidents the threshold was 1.141. Smaller accelerations cannot be excluded, but one of the magnitude that national warnings imply would have been visible.

### Control series and sensitivity analyses

The control series behave differently (Table 4). Among provider breaches that were not hacking incidents, the share affecting fewer than 5,000 individuals rose in the same direction but more weakly (1.198, 0.994 to 1.444, P = .06); among hacking incidents at entities that are not providers there was no change (0.930, 0.710 to 1.217, P = .60). The shift in victim size is therefore specific to hacking incidents at health care providers, not a property of the portal.

The fall in the monthly count at December 2022 behaved the same way: large for provider hacking incidents (0.601, P < .001) but absent in both controls (0.865, P = .43; 1.021, P = .89). A general change in how reports were received or posted would have moved all three series.

The reversal did not depend on the cutoff: the change in trend was 1.448 at 1,000 individuals, 1.258 at 2,500, 1.409 at 5,000 and 1.309 at 10,000, all P < .001, with the best-fitting breakpoint in August or October 2021 at every threshold (Table 4).

Adding the number of catalog additions in the month to the model for exploitation within 7 days did not remove the flattening after December 2022; it strengthened it (change in trend 0.055, 0.020 to 0.156, P < .001, compared with 0.144 without the covariate), although the catalog’s pace was itself associated with faster recorded exploitation (odds ratio 1.442 per unit of log additions, 1.223 to 1.701). Dropping the catalog’s first 3 or 6 months, when a backlog was being worked through, left the flattening in place (0.056 and 0.237, both P < .05) but moved the estimated pre-break rise between 3.3 and 14.0, so the size of that rise should not be read closely.

The reversal was not produced by reports still being processed: ending the series in June 2025 or December 2024, leaving 17 and 23 months for reports to accumulate, gave 1.333 and 1.374, both P < .001. Residual autocorrelation was not detectable (Ljung–Box P = .28, .09 and .26 at 6, 12 and 18 lags), and the estimates were unchanged when the Newey–West lag was set to 6, 12 or 18 (supplementary material).

## Discussion

Two changes in the threat facing US health care providers are well established in these data: the warning period between public disclosure of a vulnerability and its exploitation has settled at about a week, and hacking breaches have shifted toward smaller organizations. Neither change is attributable to generative artificial intelligence. Both began before it was widely available, neither accelerated afterward, and for breach size the best-fitting turning point was more than a year earlier.

### The warning period stopped shrinking rather than shrinking faster

Of the three series we examined, this is the one whose turning point cannot be dated before generative artificial intelligence: December 2022 sits inside the confidence set for the break. The direction, however, is the opposite of what the accelerant hypothesis predicts. The share of vulnerabilities exploited within a week had been climbing; after late 2022 it stopped climbing and timelines were stable from 2023 to 2025. This fits other reports that median exploitation times have been in single digits since 2021,^10^ and that the steep fall in average time to exploit in 2023 reflected exploitation of previously unknown vulnerabilities.^9^ One threat intelligence provider has judged that 2025 was not a year in which artificial intelligence directly caused breaches.^16^

That does not mean artificial intelligence is irrelevant. Large language models now write phishing emails that work as well as expert-written ones;^17^ health care staff already click on about 1 in 7 simulated phishing emails;^18^ and artificial intelligence-assisted extortion of health care organizations has been reported.^19^ Even so, the share of breaches listing email as a location fell from 53.8% in 2019 to about a quarter from 2023 onward, and when breakpoints were searched for that series the best fit was July 2020, long before generative artificial intelligence; in Japan phishing email accounted for only 2.2% of ransomware initial access in 2025.^20^ Greater capability has not yet appeared as faster exploitation or as a shift toward email-borne intrusion. The absence is informative rather than inconclusive: an acceleration of 21% per year or more in reported incidents would have been detected, and the point estimate runs the other way. What artificial intelligence may change is the cost of attacks rather than their timing, and that would show first in the number of attempts.

### A reversal in who is attacked

The falling share of small breaches through 2022 matches earlier findings that ransomware concentrated on large organizations between 2016 and 2021.^7,8^ The rise since then indicates that the pattern has turned; a formal test locates the turn in late 2021, it survives every size cutoff we tried, and it does not appear in either control series, which makes a change in reporting an unlikely explanation. Japan saw a parallel change: small and medium-sized enterprises made up 52% of ransomware victims in 2023 and 63% in 2024 and 2025.^20^ Another analysis of the same US breach portal found that ransomware’s share of breach counts fell after 2021 while its share of individuals affected rose,^21^ which is consistent with a split into a few very large events and many smaller ones.

The economics of ransomware offer an explanation. Initial access is opportunistic: attackers scan for devices with known weaknesses, and brokers sell that access, ranking victims by size and value.^22^ Entry therefore depends on weakness, while monetization favors larger targets. As access becomes cheaper and the hand-off between criminal groups faster,^16^ poorly defended small organizations are more likely to be compromised. Scans of German hospitals found that larger hospitals exposed more vulnerabilities, though some small facilities also exposed many.^23^ The growing involvement of business associates and other third parties in health care breaches points to vendor connections as a common way in,^24–26^ a pattern also seen in the Japanese hospital incidents for which an entry route has been disclosed publicly.^27–30^

The drop in the monthly count at December 2022 deserves comment. We had expected it to reflect a change in how reports were received or posted, but the control series rule that out: neither other breach types reported by the same providers nor hacking incidents reported by other kinds of entity show any level change at that date. Whatever produced it was specific to hacking incidents at health care providers. It is unlikely to be a fall in attacks, since annual counts resumed rising immediately afterward. We report it for completeness without being able to explain it; a one-off level shift does not affect the trend estimates on which our conclusions rest.

### Implications for critical infrastructure protection

Health care organizations take a median of 58 days to resolve serious vulnerabilities,^31^ and across sectors half of known exploited vulnerabilities remain open for months.^32^ When exploitation follows disclosure within about a week, and a quarter of vulnerabilities are exploited by the day they are disclosed, patching speed alone cannot protect most providers, and least of all small ones without security staff. A defense that requires an organization to win a race it enters two months late is not a defense.

Priorities therefore need to shift from speed to exposure and endurance. First, what is reachable from the internet should be reduced, above all the always-on remote maintenance connections opened for vendors; the share of health care breaches involving third parties makes these a standing entry route rather than an edge case. Second, core clinical systems should be segmented and access kept to the minimum needed, in line with zero-trust principles,^33^ so that one compromised account or appliance does not reach the whole estate. Third, organizations should be able to keep caring for patients without electronic systems for a period measured in weeks rather than hours, and should exercise that capability. These steps follow the health industry cybersecurity practices published by the US Department of Health and Human Services,^34^ but our findings change which organizations most need help adopting them, and argue for treating inventory of the externally reachable attack surface as a standing preparedness measure rather than a periodic audit.

The shift toward smaller organizations has a consequence that reaches beyond the organizations themselves. Because an attack on one hospital measurably degrades emergency care at its neighbors,^3–5^ a small, under-resourced provider is not only its own risk; it is a regional one. That is a standard argument for public investment, and it applies here directly: support, funding, technical assistance and reporting requirements built around large health systems will increasingly miss the organizations now most exposed, while the harm from their compromise falls on the surrounding community. Shared threat intelligence has been proposed as one way to extend capability to organizations that cannot build it alone,^35^ and it is most valuable precisely where staff are fewest. Sector-wide monitoring of where attacks land, by organization size, would make such shifts visible early enough to act on; the news-derived mapping of the threat landscape proposed elsewhere^13^ and the reporting data used here are complementary inputs to such monitoring.

### Limitations

Catalog addition dates are not the dates of first exploitation, and the catalog is not exhaustive. The vulnerability series has only 13 months before the break, so those estimates are less precise than the breach estimates, the magnitude of the pre-break rise is sensitive to how much of the catalog’s first year is included, and the break in that series cannot be dated relative to the arrival of generative artificial intelligence. The number of individuals affected is an imperfect proxy for organization size; breaches affecting fewer than 500 individuals are not reported, and the breach classifications in this source contain errors.^36^ Reported breaches may not represent all attacks, and remediation times come from industry sources that are not peer reviewed. Interrupted time-series analysis identifies changes in level and trend around a date; it does not establish cause, and other events in the same period, including large third-party incidents, could produce similar patterns. Our conclusion about generative artificial intelligence is therefore a negative one — we find no acceleration coinciding with it, and can exclude accelerations above the bounds reported — rather than a claim about what did cause the changes. The control series strengthen the case that the shift in victim size is real rather than an artifact of reporting, but they cannot identify its cause either.

## Conclusions

By the time generative artificial intelligence became widely available, the warning period between disclosure of a vulnerability and its exploitation had narrowed to about a week; it then stopped narrowing, and we cannot date that turn to before or after the technology arrived. Breaches at US health care providers have shifted toward smaller organizations, a reversal that a formal test dates to late 2021 and that does not appear in control series from the same reporting system. Neither change is an acceleration, and the growth in reported incidents did not speed up after generative artificial intelligence arrived. Preparing the health sector for this threat means reducing what is exposed and sustaining care when systems fail, and directing national support toward the smaller organizations that are now bearing the attacks and whose failure is felt across their regions.

## Supporting information

Data and code archive

## Acknowledgements

The authors used Claude (Anthropic; Opus 5.5 and Opus 5) as a writing and programming aid in preparing this work: for language editing of the manuscript, for implementing in Python the statistical analysis and the figures specified by the authors, and for formatting the bibliography. The research question, the study design, the choice of models and the interpretation of the results are the authors’ own. The authors reviewed the analysis code, re-ran it, and checked every number and figure reported here against the analysis output. The authors have reviewed and edited all AI-assisted content and take full responsibility for the published work.

## Author contributions

Ryutaro Nakagawa: conceptualization, methodology, formal analysis, writing – original draft, writing – review and editing. Yuya Yasuhara, Akiyo Shimoyama, Yuki Nishi and Yuki Kubota: data curation, formal analysis, writing – review and editing. All authors approved the final manuscript.

## Ethics

The study used only publicly available aggregate or organization-level records. No individual-level health information was accessed and no human participants were involved, so institutional review board approval was not required.

## Data availability

All source data are public: the Known Exploited Vulnerabilities catalog of the US Cybersecurity and Infrastructure Security Agency, the CVE List V5 repository, and the breach portal of the Office for Civil Rights of the US Department of Health and Human Services. The processed monthly, annual and stratified series, the full regression and robustness output, and the analysis code are attached to this preprint as a supplementary file, under a Creative Commons Attribution licence for the data and an MIT licence for the code, so that every analysis reported here can be reproduced.

